# Confirmation of multi-parallel quantitative real-time PCR as the gold standard for detecting soil-transmitted helminths in stool

**DOI:** 10.1101/2021.12.09.21267271

**Authors:** Marina Papaiakovou, Nils Pilotte, Julia Dunn, David TJ Littlewood, Rubén O Cimino, Alejandro Krolewiecki, Steven A Williams, Rojelio Mejia

**Affiliations:** Natural History Museum, Cromwell Road, SW7 5BD, London, UK; London Centre for Neglected Tropical Disease Research, Imperial College London, London, W2 1PG, UK; Department of Biological Sciences, Smith College, Northampton, MA 01060, USA; Clinton Health Access Initiative, Lao PDR; Instituto de Investigaciones de Enfermedades Tropicales (IIET-CONICET), Sede Regional Orán, Universidad Nacional de Salta, Argentina; Department of Pediatrics, Section of Tropical Medicine, National School of Tropical Medicine, Texas Children’s Hospital and Baylor College of Medicine, Houston, TX 77030, USA

## Abstract

Due to its simplicity and cost-effectiveness, microscopy has seen extensive field-use as the diagnostic standard for the detection of soil-transmitted helminths (STH) in stool samples. However, the sensitivity of microscopy-based detection is inadequate in reduced-transmission settings where worm burden is oftentimes low. Equally problematic, eggs of closely related species oftentimes have indistinguishable morphologies, leading to species misidentification. In light of these shortcomings, the purpose of this study was to demonstrate multi-parallel quantitative real-time PCR (qPCR) as the new “gold standard” for STH detection. Accordingly, stool samples from non-endemic participants were spiked with limited numbers of eggs or larvae (1 to 40) of five different species of STH. DNA extracts were tested using two unique multi-parallel real-time PCR-based diagnostic methods. These methods employed different target sequences (ribosomal internal transcribed spacer, or highly repetitive non-coding regions), to evaluate the detection of DNA from as little as one egg per sample. There was a statistically significant kendall correlation between egg/larvae counts and qPCR from both methods for Trichuris trichiura (0.86 and 0.872 for NHM and Baylor assays) and a strong correlation (0.602 and 0.631 for NHM and Baylor assays, respectively) for Ascaris lumbricoides. Less strong but still significant was the Kendall Tau-b value for A. duodenale (0.408 for both) and for S. stercoralis (0.483 and 0.653, respectively). In addition, using field stool samples from rural Argentina both assays had fair to moderate kappa agreement (0.329-0.454), except for Strongyloides stercoralis (0.121) that both assays had slight agreement. In spite of the small cohort of samples, both qPCR assays, targeting of two independent genomic regions, provided reproducible results and we believe that, low cost multi-parallel quantitative real-time PCR-based diagnostics should supplant microscopy as the new gold standard for stool-based detection of soil transmitted helminths in public-health and community settings.

## Introduction

Soil transmitted helminths (STH) infect over 1.6 billion people worldwide,^1^ resulting in years of disability and extensive morbidity. The most widely employed techniques for diagnosing STH infections remains microscopy-based, despite the repeated demonstration of their shortcomings. ^2–6^ The use of such techniques hinders efforts aimed at the accurate assessment of infection prevalence and worm burden due to the poor sensitivity and specificity of the tools, particularly when infection levels are low at the community level. ^7–9^ Despite the repeated demonstration of the limitations of microscopy-based methods^2^ they remain the World Health Organization (WHO)-recommended techniques for use in parasitological and epidemiological interventions and are almost universally used for the monitoring and evaluation of deworming programmes. ^10^ However, the new WHO roadmap 2021-2030 still supports the development and implementation of a more sensitive tool to confirm the cessation of an MDA programme and the success of an intervention. ^11^

Over the last two decades, the accretive development of more sensitive molecular tools for the diagnosis of protozoan and bacterial infections have spurred interest in the similar development of tools for monitoring STH infections. ^11–14^ These advances in molecular testing have largely utilized real-time polymerase chain reaction (PCR). The primary advantage of real-time PCR over microscopic techniques is its capacity to detect DNA from shed parasite material, even when infections are of low intensity, or prevalence is minor. Under such conditions, eggs are more easily missed by microscopy. Moreover, real-time PCR does not require the use of fresh specimens, a requirement for the microscopic detection of certain helminths including various hookworm species and *Strongyloides stercoralis*. Real-time PCR also allows for the parallel detection of other parasitic agents (e.g. other helminths such as *Schistosoma*, or protists) and the DNA extraction procedures required for such assays typically do not preclude the conducting of microbiome studies from the same purified extracts. ^15–18^ Utilizing PCR the capacity to harmonize testing across labs also increases, particularly when automation is involved. ^11,19,20^ In contrast, microscopic methods are heavily reliant upon extensive training, and operator skill and ability, making inter-lab consistency less easily attainable. Also, real-time PCR is quantitative and can report the amount of parasite target DNA in a stool sample, while certain microscopy techniques (e.g. McMasters, Kato-Katz) is semiquantitative.^21,22^ Lastly, and importantly, species appearing to be identical under the microscope can be differentiated by real-time PCR. However, despite these advantages, the labour– and equipment–associated costs of both stool processing and the subsequent molecular analyses have raised concerns over the feasibility of the technique’s routine use. To overcome these obstacles, efforts are underway to simplify such techniques, making the underlying technologies increasingly practical, cost-effective, and field-deployable, in turn facilitating the use of improved diagnostics in the most challenging of settings. ^11,13,14,23^

Recognizing that intervention success and informed decision making is highly reliant upon the use of sensitive and accurate diagnostic tools, the incorporation of molecular methodologies into various programmatic and research efforts. ^11,24–28^ Such increased use has resulted in several attempts to redefine the ‘gold standard’ for STH detection. However, attempts to comparatively evaluate molecular methods remain minimal, and nearly all published evaluations compare one or more microscopic techniques with a solitary molecular method. ^8,29–33^

Distinct real-time PCR assays targeting a variety of DNA regions (ribosomal internal transcribed spacer sequences [ITS], ribosomal subunit sequences, or mitochondrial genes) have been developed in recent years for the detection of STH^12^. Assays that target ribosomal or mitochondrial sequences can be found in adequate copy number, providing moderate to high sensitivity, making them attractive real-time PCR targets. However, while attractive, these targets remain suboptimal. The development of new bioinformatics tools has significantly facilitated the identification and optimization of assays that target highly repetitive elements, reducing both the time and money expended for their identification. Such targets have improved both the sensitivity and specificity of real-time PCR assays, enabling the differentiation of closely related species, and facilitating target detection at copy numbers below those found within a single egg.^11,34–36^

The goal of this study was to provide an assessment of the overall agreement when two distinct molecular assays utilizing different target sequences are compared across a panel of samples spiked with known quantities of STH eggs or larvae. Similar testing was also performed on a panel of field-collected samples, to assess the transferability of results. General agreement would provide strong evidence for the definitive consideration of real-time PCR as the gold standard for STH detection. Results will also highlight platform-specific strengths and weaknesses, informing choices in an application-specific manner. To the best of our knowledge, this is the first comparison of two independent real-time PCR platforms, with unique molecular targets, utilizing samples spiked with known numbers of STH eggs or larvae.

## Materials and methods

### Ethical Approval, consents and permissions

This project was approved by the bioethics committee of Colegio de Médicos de la Provincia de Salta on 19 March 2015 and the internal review boards of Baylor Medical College of Medicine (protocol number H-34926). Informed written consent was obtained from each subject or from subject’s parent/guardian as appropriate. All patients in Orán, Argentina were treated, free of charge, with albendazole and ivermectin (prior to the study or after based on microscopy findings.

### Field Sample Collection

A panel of 130 samples was collected from a random cohort of subjects ages five to 12 years old living in Salta, Argentina. Collection of samples were part of ongoing field studies. All samples immediately underwent microscopic analysis and were subsequently placed in the freezer, without preservative, until DNA extraction and subsequent qPCR analysis occurred.

### Microscopy

Stool samples were examined by Telemann concentration microcopy, Baermann, Harada Mori, McMaster egg-counting technique, with some modifications^8^ and sedimentation method at the time of sample drop-off, using protocols previously published.^8^ The McMaster method was performed following a standard procedure: 2g of stool was filtered and homogenized with 30ml of saturated saline solution. Upon loading two flotation chambers for each sample, the eggs were allowed to float for five minutes and then counted. The number derived, was multiplied by 50, in order to calculate eggs per gram (epg).

Real-time PCR was compared directly with samples that were positive with any of the microscopy techniques employed.

### Control stool for spiking

The control human stool used in the study was collected from an individual in a non-endemic area for parasites who had never travelled. Stool was immediately stored at - 20 °C. At the time of collection.

### Parasite/Egg collection for spiking

Larvae and STH eggs were collected from a field clinic in Orán, Argentina. Upon arrival at the clinic, fresh fecal material underwent processing for the identification of STH and protozoa present in the specimen. All processing occurred using the McMaster’s technique (magnification). Eggs/larvae present within positive samples were carefully removed from the samples under the microscope, rinsed with nuclease free water, and placed in cryovials for storage at −20 °C. Samples remained at −20 °C until shipment to Baylor College of Medicine where known quantities of eggs/larvae were utilized for the spiking of the control stool samples.

### Spiked stool & DNA extraction

Known numbers of parasitic eggs/larvae (1, 2, 5, 10, 20, 40) were used to spike 10 mg samples of naïve stool, at Baylor College. The entire sample then underwent DNA extraction using the Fast DNA Spin Kit for Soil (MP Biomedicals, Santa Ana, CA) as previously described.^7^ One half of the elution volume of each sample was kept at Baylor College for qPCR testing, and the remaining volume was shipped to the Natural History Museum for parallel independent analysis.

### Field samples & DNA extraction

A total of 500 mg of stool from each field sample was subjected to DNA extraction at Baylor College of Medicine. Samples were again extracted using the Fast DNA Spin Kit for Soil as described above. However, to facilitate the detection of *T. trichiura*, an additional step was incorporated into the procedure. This addition to the protocol resulted in a second elution volume from each sample, specifically for use with the *T. trichiura* detection assay.^7^ As described above, one half of the elution volume from each sample remained at Baylor College of Medicine where all samples underwent qPCR analysis, while remaining aliquots were sent to Smith College for parallel and independent testing.

### Multi-parallel real-time PCR assays

DNA extracts from all spiked samples were assayed independently at both Baylor College of Medicine and the Natural History Museum (NHM).

#### ITS regions and 18S rRNA-targeting real-time PCR (Baylor College)

At Baylor College of Medicine, multi-parallel qPCR was performed using the same reaction conditions and employing the same species-specific (except for *Ancylostoma*) primers and FAM-labeled minor-groove-binding probes as previously described^7^ (ThermoFisher, Waltham, MA). Henceforth, these assays will be referred to as ‘Baylor assay’. All samples were run on a ThermoFisher ABI ViiA7 PCR machine in duplicate with default parameters and 40 cycles as the cut-off. DNA concentrations were calculated using a standard curve.

#### Repeat-targeting real-time PCR (Smith College/NHM)

At the Natural History Museum, multi-parallel qPCR was performed using the same reaction conditions and employing the same species-specific primers and FAM-labelled double quenched probes as previously described. ^34^ Hereafter, these assays will be referred to as ‘NHM assay’. All samples were tested using a StepOne Plus Real-time PCR instrument and all reactions were run in duplicate. For each sample, DNA target copy number per ul of reaction was calculated based on an intra-plate standard curve (run in triplicate).

### Statistical analysis

To assess the correlation between DNA quantity (fg/ul or copies/ul) and spiked egg numbers, the Kendall rank correlation test was used (preferred for small datasets). The Kendall Tau-b value was chosen as it adjusts for tied ranks. Kendall Tau-b values range between minus one (all pairs are discordant) and one (all pairs are concordant); a higher Tau-b value indicates more concordant than discordant pairs of individual egg counts and therefore higher overall correlation^37^; interpretation as < + or - 0.10: very weak; + or −0.10 to 0.19: weak; + or - 0.20 to 0.29: moderate; + or - 0.30 or above: strong. The correlations were visualized in R version (3.1.3); *Necator americanus* was excluded from the graphs due to few data points, but the correlation values are still shown in Table 1.

**Table 1.** Kendall correlation as Tau-b value and respective p values, between number of larvae/eggs and qPCR quantitative method for each soil-transmitted helminth for both NHM/Smith and Baylor assays

| Species | Kendall correlation egg/larvae vs quantitative method |  |  |  |
| --- | --- | --- | --- | --- |
|  | NHM | p-value | Baylor | p-value |
| *Ancylostoma duodenale* | 0.408 | 0.44 | 0.408 | 0.44 |
| *Ascaris lumbricoides* | 0.602 | 0.0022 | 0.631 | 0.0021 |
| *Necator americanus* | 0 | 1 | -0.816 | 0.12 |
| *Strongyloides stercoralis* | 0.483 | 0.0022 | 0.653 | 2.30E-05 |
| *Trichuris trichiura* | 0.86 | 6.30E-07 | 0.872 | 4.50E-07 |

P values <0.05 were considered statistically significant. The comparisons between the two distinct molecular approaches were depicted using unweighted Cohen’s Kappa agreement ^38^ which accounts for the possibility that concordance between the two assays compared herein may occur by chance. Kappa < 0 means ‘No agreement’; 0 — .20 defines ‘slight agreement’; .21 — .40 ‘Fair’; .41 — .60 ‘Moderate’; .61 — .80 ‘Substantial’; .81–1.0 ‘Perfect’. Fleiss kappa was calculated to evaluate the agreement between the microscopy and the 2 qPCR tests (3 raters), treating the results as categorical values (presence/absence). All kappa agreements were calculated using R version 3.1.3 and R package ‘irr’.

## Results

### Spiked samples

The Kendall Tau-b value was 0.86 and 0.872 for *T. trichiura*, for both NHM and Baylor assays, respectively, indicating a strong concordance between DNA quantity measured using qPCR and egg numbers. For *A. lumbricoides*, the Tau-b value of 0.602 and 0.631 for NHM and Baylor assay, respectively, also showing a strong concordance. Less strong but still significant was the Tau-b value for *A. duodenale* (0.408 for both), using both assays, and for *S. stercoralis* (0.483 and 0.653, respectively). Results for *N. americanus* remain puzzling (with 0 and −0.816 Taub-b value for NHM and Baylor, respectively), probably due to insufficient number of eggs/larvae for that species (Table 1).

The graphs in Figures 1 and 2 also show the linearity and correlation between the qPCR quantitative method and eggs or larvae spiked.

**Figure 1.**
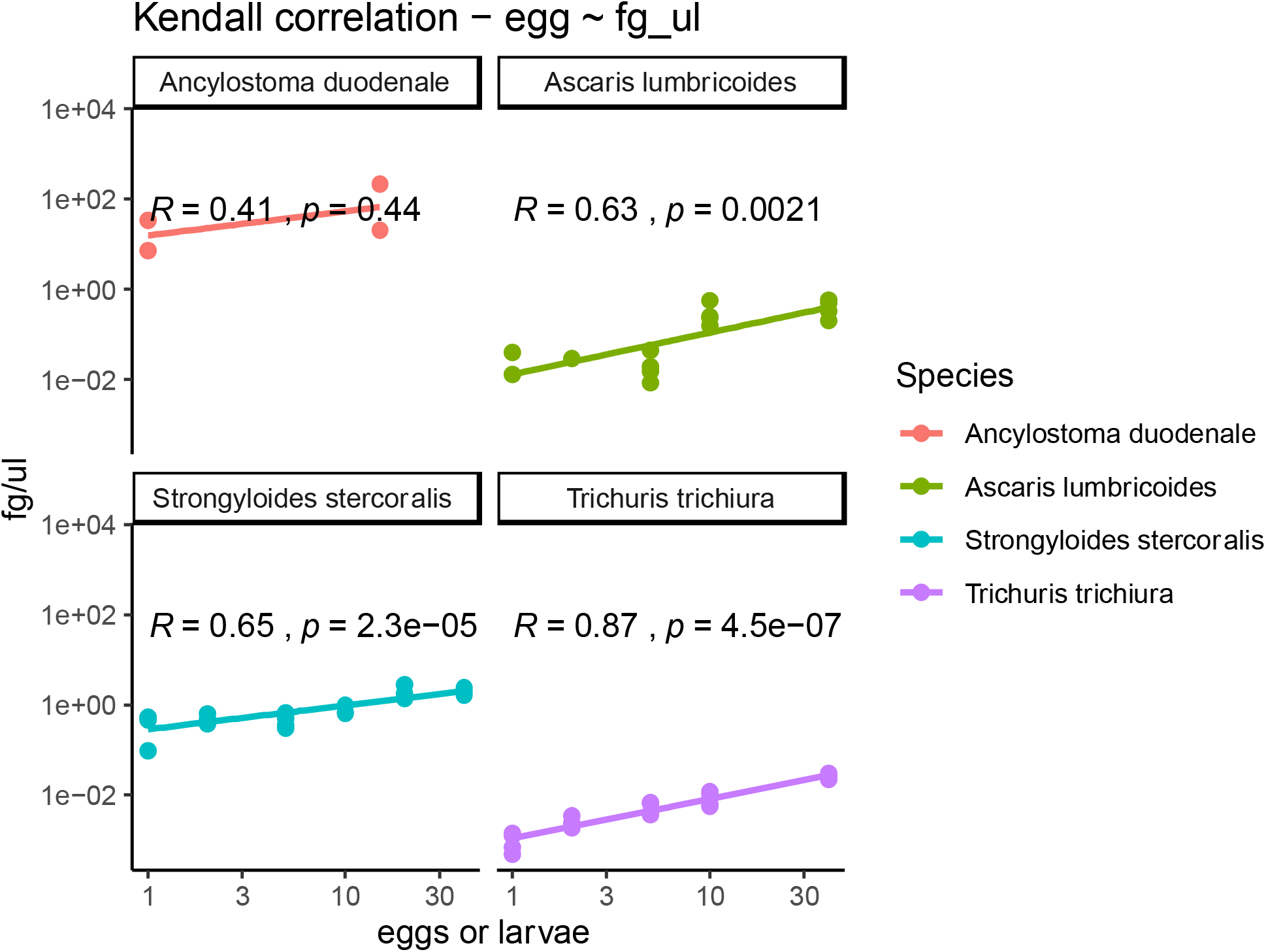
Kendall correlations for Baylor assays, between eggs or larvae and fg/ul for all soil-transmitted helminths except for N. americanus due to in insufficient number of larvae

**Figure 2.**
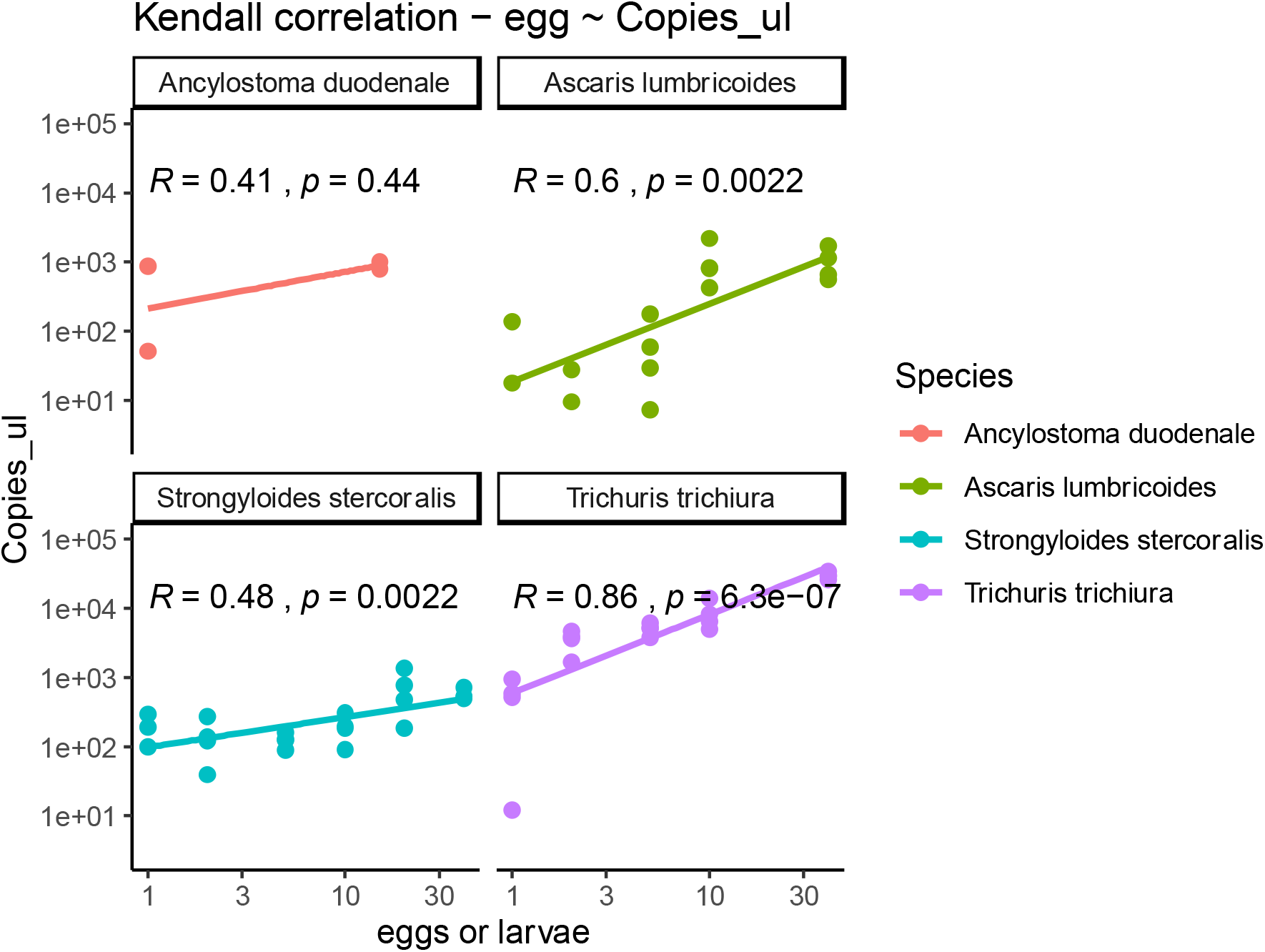
Kendall correlations for NHM/Smith assays, between eggs or larvae and fg/ul for all soil-transmitted helminths except for N. americanus due to in insufficient number of larvae

### Field samples

We calculated both overall percentage agreement (total number of agreed positives or negatives in a given sample set) and Cohen’s kappa for agreement on a sample-by-sample case, for both sets of qPCR assays, treating data as categorical values (presence/absence), since the qPCR output interpretation is still not corresponding directly to worm burden or worm intensity. To show the greater discordance between microscopy and both qPCR assays, we calculated Fleiss kappa; results are show in Table 2.

**Table 2.** Cohen’s kappa (unweighted) for inter-rater reliability comparison between the two qPCR methods, treating the data as categorical variables (presence/absence); kappa cheat sheet *<0 No agreement; 0 — 0*.*20 Slight; 0*.*21 — 0*.*40 Fair; 0*.*41 — 0*.*60 Moderate; 0*.*61 — 0*.*80 Substantial; 0*.*81–1*.*0 Perfect*

|  | Two qPCRs - RJ and NHM (two raters) |  |  | Fleiss kappa |  |
| --- | --- | --- | --- | --- | --- |
|  | Unweighted Cohen's Kappa agreement | Percentage agreement | p-value | Fleiss kappa (categorical) | p-value |
| *Necator americanus* | 0.329 | 82% | 6.66E-16 | 0.214 | 0 |
| *Trichuris trichiura* | 0.366 | 96% | 0 | 0.178 | 1.53E-09 |
| *Ancylostoma duodenale* | 0.279 | 64% | 3.77E-15 | 0.152 | 6.39E-11 |
| *Strongyloides stercoralis* | 0.121 | 77% | 0.000471 | 0.0639 | 0.0186 |
| *Ascaris lumbricoides* | 0.454 | 92% | 0.00E+00 | 0.219 | 9.14E-12 |

Between the two sets of qPCR assays, there was a moderate agreement for A. lumbricoides and a fair agreement for *N. americanus (kappa value = 0*.*329), T. trichiura (kappa value = 0*.*366)* and *A. duodenale (kappa value = 0*.*279)*, though both assays showed a slight agreement for *S. stercoralis (kappa value = 0*.*121)*.

Fleiss kappa showed slight agreement between microscopy (ranging from 0.0639 to 0.219) and both qPCR assays (Table 2).

## Discussion

We present a comparative study evaluating two independent, quantitative real-time PCR assays for five soil-transmitted helminths using both spiked and field samples. By comparing spiked samples using different target DNA sites, we showed concordance and moderate to strong correlation not only to the presence of helminth egg or larvae, but also quantitatively to the amount of parasite DNA.

Such strong correlation between spiked eggs and qPCR output has been previously demonstrated in similar settings. ^33^ However, this is the first pilot study utilizing two sets of qPCR targeting different genomic sequences.

The great discordance between both qPCR sets and microscopy depicts the superiority in sensitivity (true positives) and specificity (true negatives) of the molecular methods compared to coprological tests.

### 1. qPCR a potential tool for “true negatives”

A key advantage of using qPCR is the high negative predictive value of determining a sample that does not have a parasitic infection. ^8^ The value of a test with high negative predictive value, such as qPCR, is evident in public health and epidemiology endeavors. Population studies on the community prevalence of STH relies on the number of subjects that do not have a parasite. Thus, demonstrating not needing therapy or follow up care provides data for planning and prioritizing resource for individual care. The ability to rule out a STH diagnosis is also valuable in mass drug administration (MDA) programs. A test that can reliably show the efficacy of MDA on a population level will note progress of STH eradication campaigns.

### 2. Monitoring the qPCR outputs, pre and post MDA

Another value, especially in MDA programs, is monitoring the change of parasitic DNA post-treatment. By using these molecular assays, the quantity of parasite DNA, which is a proxy for parasite eggs and larvae, can be observed and followed for treatment efficacy or failure. Since the two tests presented in this study detect different DNA sequences, they cannot be used interchangeably when monitoring changes in DNA concentrations. Similar studies to the current one have shown qPCR as a promising tool to estimate intensity of infection ^29,33^ but further studies are still needed to not only identify a universal consensus for qPCR diagnostics for STHs, but to also to establish qPCR as a standalone tool for estimating worm burden in the absence of coprological methods.

Thus, whilst still more studies are needed to achieve a reliable correlation between epg number and intensity of infection to qPCR outputs, a near term goal moving forward would be to collectively assess the parasitic burden through PCR output, in a population as a whole, rather than at the individual level.

### 3. Capacity building

An important component of using these molecular assays is transferring the technology to build capacity in resource-limited areas. ^8,29,32,39,40^ Supporting the WHO 2021-2030 roadmap^41^, building diagnostics facilities in-country can prove beneficial beyond diagnostics for STHs. Similar workflows can be used for screening of other parasites and knowledge transfer on diagnostics can create empowerment in local, in-country groups and generate more research funds and establish capacity building and integrate training as part of routine health activities.

### 4. Limitations

We acknowledge the number of spiked samples (eggs and larvae) trialed in this study was limited and requires further scaling. qPCR could replace coprological tools in low-intensity endemic for STHs area, therefore, we only focused on spiking stool samples with very low numbers of eggs/larvae. Another limitation was that an extraction control, to check the efficacy of the DNA extraction method, was not available. Perhaps a ‘failed’ extraction in the samples spiked with a single egg explains the 0 and negative correlation for both assays for *N. americanus*. Controls should be used, not only for accurate quantitation of PCR via standards, but for both the efficacy of the extractions and the efficacy of the PCR (i.e., detecting inhibition).

### 5. Conclusion

We present supporting evidence that qPCR is a valid replacement for fecal microscopy and our brief, pilot exercise supports current work ^33^ towards replacing coprological tools to assess low STH infections using PCR. Different laboratories prefer and favor different targets, but our pilot study has shown that any PCR is better than no PCR at all when STH infections are low in a population. Using spiked and field samples we conclude that qPCR can be pursued towards a gold standard for detection of STH and can be successfully deployed in resource-limited countries.

## Data Availability

All data produced in the present study are available upon reasonable request to the authors.

